# COVID-19 in people living with human immunodeficiency virus: A case series of 33 patients

**DOI:** 10.1101/2020.04.28.20073767

**Authors:** Georg Härter, Christoph D. Spinner, Julia Roider, Markus Bickel, Ivanka Krznaric, Stephan Grunwald, Farhad Schabaz, Daniel Gillor, Nils Postel, Matthias C. Mueller, Markus Müller, Katja Römer, Knud Schewe, Christian Hoffmann

**Author notes:** **Correspondence:** Georg Härter, MD, Medicover MVZ Ulm, Münsterplatz 6, D-89073 Ulm, Germany. **Conflict of interest**: The authors have declared no conflicts of interest.

## Abstract

Data on people living with human immunodeficiency virus (PLWH) in the current SARS-CoV-2 pandemic is still scarce. This case series of 33 PLWH patients with COVID-19 reveals symptoms and outcome in this special population. Three out of 32 patients with documented outcome died (9%). However, 91% of the patients recovered and 76% have been classified as mild cases, indicating that there is no excess morbidity and mortality among PLWH with symptomatic COVID-19. All patients were on antiretroviral treatment, of them 22 on tenofovir-containing regimen, and 4 on the protease inhibitor darunavir.

## Introduction

On April 27, the number of confirmed worldwide SARS-CoV-2 infections has exceeded up to nearly 3 million cases and almost 200,000 deaths [1]. Consequently, increasing numbers of coronavirus disease (COVID-19) cases are expected to rapidly occur in people living with human immunodeficiency virus (PLWH). In the current COVID-19 pandemic, several comorbidities have been identified as risk factors for severe disease and death [2–5]. Data on PLWH is still scarce. A small case series from Spain recently described the clinical characteristics of five PLWH with COVID-19 [6]. Coronaviruses such as severe acute respiratory syndrome (SARS)-CoV and SARS-CoV-2 have been shown to cause transient immune deficiency [7–9], indicating that HIV and COVID-19 may both carry deleterious immunological and clinical consequences. On the other hand, defective cellular immunity in PLWH could paradoxically be protective for severe cytokine dysregulation, which has been observed in patients with COVID-19. Moreover, some HIV protease inhibitors (PI) are thought to inhibit the 3-chymotrypsin-like protease of coronaviruses.

In this study, we describe our early experiences with COVID-19 and clinical characteristics in patients with documented HIV infection.

## Methods

This retrospective analysis included all cases of PLWH with SARS-CoV-2 infection, which were confirmed between March 11 and April 17, 2020 in 12 participating German HIV centers. Anonymized data were collected by the treating physicians and included age, gender and HIV-associated parameters such as the last CD4+ T-cells (absolute cells/mm^3^, as assessed by local labs), the last CD4/CD8 Ratio, the last HIV-RNA (copies/mL, as assessed by local labs) and antiretroviral therapy before COVID-19 diagnosis. With regard to COVID-19, clinical symptoms, severity of disease classified as mild (i.e. non-pneumonia and mild pneumonia), severe (i.e. dyspnea, respiratory frequency ≥ 30/min, blood oxygen saturation ≤ 93%, and/or lung infiltrates > 50% within 24 to 48 hours), and critical (i.e. respiratory failure, septic shock, and/or multiple organ dysfunction or failure) [10], and outcome were collected as well as comorbidities. Ethics committee approval was obtained from Technical University of Munich (April 4, 2020, Approval No. 194/20s).

## Results

We identified 33 PLWH with confirmed SARS-CoV-2 infection. Positive SARS-CoV-2 PCR was obtained from nasopharyngeal swabs in 29, and from bronchoalveolar lavage or sputum in 2 cases, in two cases no information about this was available. For 14 patients a close contact to a person with SARS-CoV-2 infection have been documented. For seven patients a travel history to foreign countries with a high transmission rate of SARS-CoV-2 have been reported. 26 patients were primarily diagnosed in the outpatient setting. In 7 patients the diagnostic procedure was done in the hospitals during admission. Additionally, neither clusters of transmission nor nosocomial infections could be detected. Main characteristics are shown in **Table 1**. Mean age was 48 years (range, 26 to 82 years) and 30/33 patients were male. All patients were on antiretroviral therapy at the time of COVID-19 diagnosis. Antiretroviral regimens included nucleoside reverse transcriptase inhibitors (NRTIs) in 31, integrase strand transfer inhibitors (INSTI) in 20, protease inhibitors (PI) in 4 and Non-NRTIs in 9 cases. NRTIs were mainly tenofovir alafenamide (16 cases), tenofovir disoproxilfumarate (6 cases) and a cytidine analogue, either emtricitabine (n=22) or lamivudine (n=9). The last median CD4+ T-cell count before SARS-CoV-2 infection was 670/mm^3^ (range, 69/mm^3^ to 1715/mm^3^). In 30/32 cases, the last HIV-RNA was below 50 copies/mL. Two patients with detectable HI-viremia needed hospital admission including intensive care treatment and mechanical ventilation, and one of these patients died. Comorbidities other than HIV infection were documented in 20/33 (60%) patients, including arterial hypertension (n=10), chronic obstructive pulmonary disease (n=6), diabetes mellitus (n=4), cardiovascular disease (n=3) and renal insufficiency (n=2). Coinfection with hepatitis B have been documented in five patients: a resolved hepatitis B (hepatitis B surface-Antigen negative) in four patients, and in one patient a chronic hepatitis B (hepatitis B envelope Antigen positive). In one patient, a cured hepatitis C (sustained virologic response after 12 weeks of treatment with sofosbuvir / velpatasvir) has been reported.

The most common symptoms were cough in 25/32 (78%), fever in 22/32 (69%), arthralgia/myalgia 7/32 (22%), headache 7/32 (22%), and sore throat in 7/32 (22%). Sinusitis and anosmia occurred in 6/32 (19%) for each. At the last available follow up, 29/32 of patients with documented outcome (91%) had recovered from COVID-19. Altogether 14/33 (42%) patients were admitted to hospitals. Treatment on intensive care units (ICU) was necessary in 6 of 14 (43%) hospitalized patients. Of the 14 patients, requiring treatment in hospitals, 10 have been discharged in the meanwhile. One patient is still in hospital but discharged from ICU. In one patient, a spontaneous pneumothorax could be seen as a complication of persisting cough. Three out of 32 patients with documented outcome (9%) had died (patient #9 aged 82 years, patient #20 with a CD4+ T-cell count of 69/mm^3^ and a very low CD4/CD8 Ratio of 0.06, and one patient #24 with several comorbidities as hypertension, chronic obstructive pulmonary disease, and diabetes mellitus type 2). The clinical case definition was mild in 25/33 cases (76%), severe in 2/33 cases (6%), and critical in 6/33 cases (18%).

## Discussion

In the current COVID-19 pandemic, comorbidities such as arterial hypertension, cardiovascular disease, diabetes and cancer have been identified as risk factors for severe diseases [2–5]. However, as these cohort studies did not provide data on HIV infection, it remains unclear whether PLWH remain at higher risk for SARS-CoV-2 infection or at higher risk for severe courses. Previous studies on influenza viruses did not find an increased morbidity and mortality in PLWH [11, 12]. For SARS and COVID-19, a few case reports have indicated no severe courses even in AIDS patients [13, 14]. In the absence of controlled and/or larger data, a preliminary statement from the European AIDS Clinical Society explained that “there is no evidence for a higher COVID-19 infection rate or different disease course in people with HIV than in HIV-negative people” so far [15].

In the present case series on 33 patients infected with symptomatic SARS-CoV-2 infection, 29/32 (91%) have recovered at the last follow up and 76% have been classified as mild cases. However, 24% of the cases have been categorized as severe or critical cases. Three patients had died (9%). The following details provide some explanations: One patient was of older age (82 years) and had a detectable viral load before COVID-19. In the other deceased patient, only limited information was available but his last CD4 T-cell count and CD4/CD8 Ratio was very low. The third patient suffered from several comorbidities as arterial hypertension, chronic pulmonary obstructive disease, and diabetes mellitus type 2. The case fatality rate of 9% is therefore higher than in the general population in Germany, where about 5,640 patients of 154,175 confirmed COVID-19 cases died (3.7%) [16]. Additionally, the number of severe and critical cases in our cohort (24%) seems to be somewhat higher than reported from other cohorts (19%) [10]. The hospitalisation rate in our cohort was 42% and therefore higher than in the general population in Germany, where the hospitalisation rate of COVID-19 patients is about 17% [16]. Of the 14 hospitalized patients 6/14 (43%) needed an admission to an intensive care unit. On the first look, this seems to be higher than it was reported in a large cohort in New York, where the ICU admission rate of patients with documented outcome was 14% [17]. This could be due to involvement of two large university hospitals where only hospitalized patients were included. In addition, there might be an effect of higher hospitalisation rates among patients with known HIV infection due to safety reasons. Nevertheless, the precise hospitalisation rate among symptomatic cases is not known. In our cohort, only symptomatic patients were documented. In larger cohorts, it has been estimated that about 20-40% of SARS-CoV-2 infected people are asymptomatic [18, 19].

Therefore, it is very likely that we have overestimated total morbidity and mortality. Another aspect is the different mean age in large cohorts (63 years) [17] and in our cohort. The mean age of 48 years in our cohort may correspond to the younger HIV infected population but also to the lower mean age of SARS-CoV-2 infected people in Germany of 50 years [16]. The main symptoms reported in our cohort were mainly cough and fever. The clinical characteristics of COVID-19 did not appear to differ from those of the general population [2, 5, 16].

All patients were on ART and all except four had CD4+ T-cells > 350/mm^3^, indicating no severe immune deficiency. However, two patients with a detectable viremia required mechanical ventilation. Nevertheless, the data obtained did not reflect whether this was due to an insufficient antiretroviral treatment regimen, a treatment failure, or maybe due to the present COVID-19 disease. There is some data on immunological consequences from two retrospective studies of 21 and 44 HIV negative patients with COVID-19, showing significant decreases of CD4+ T-cells in almost all patients, with a more pronounced decline in severe cases [8, 9]. There is also evidence from a larger study on SARS-CoV, showing a prolonged lymphopenia before returning towards normal after five weeks, with the lowest mean CD4+ T-cell count of 317 cells/µl [7]. Up to now, it remains unclear whether this may translate into a higher risk for opportunistic infections.

We have identified 4/33 patients who acquired SARS-CoV-2 while these patients were treated with a PI containing regimen, consisting in boosted darunavir in all cases. This percentage does not appear to differ markedly from the total PLWH population in Germany, in which the proportion of patients on boosted PIs has constantly decreased during recent years [20, 21]. For another HIV-PI, lopinavir, uncontrolled studies have indicated a potential benefit in COVID-19 with early initiation [22–24]. In addition, a case-control study on Middle East Respiratory Syndrome (MERS) has suggested an effect for lopinavir/r as post-exposure prophylaxis in health care workers [25]. However, the first randomized open-label trial in 199 adults with severe COVID-19 did not find any clinical or virological benefit with lopinavir/r beyond standard care [26]. It was speculated that concentrations of protein-unbound lopinavir achieved by current HIV dosing are too low for inhibiting viral replication. Nevertheless, several trials of lopinavir and darunavir are ongoing, including a cluster-randomized clinical trial on 3.040 participants in Spain (HCQ4COV19). Our preliminary findings did not suggest a protective effect of darunavir which is in line with US Guidelines, recommending that ART regimen “should not be changed to include a PI to prevent or treat COVID-19, except in the context of a clinical trial and in consultation with an HIV specialist” [27].

Beside PI, we did not find a clear evidence for a protective effect of tenofovir. Of note, the nucleoside analogue remdesivir, which is currently tested in several clinical trials for COVID-19 [28], has some chemical similarities to tenofovir alafenamide. In molecular docking studies, tenofovir has been recently shown to bind to SARSCoV-2 RNA polymerase (RdRp) with binding energies comparable to those of native nucleotides und to a similar extent as remdesivir. Consequently, tenofovir has recently been suggested as a potential treatment for COVID-19 [29]. In Spain, a large randomized phase 3 placebo-controlled study compares the use of tenofovir disoproxil fumarate/emtricitabine, hydroxychloroquine or the combination of both versus placebo as prophylaxis for COVID-19 in healthcare works [30]. As the majority of our patients (22/33) was treated with tenofovir alafenamide or tenofovir disoproxil fumarate, including those developing severe or critical disease, our cohort data indicate no or only minimal clinical effect of tenofovir against SARS-CoV-2.

There is no doubt that our study has important limitations. First, this was a small retrospective and uncontrolled case-series with limited follow-up. All patients were symptomatic, indicating that asymptomatic cases were probably missed. For the antiretroviral treatment regimen, we did not have the precise rates of overall PI or tenofovir prescriptions in the participating centers. Other important data were incomplete, including transmission and exposure conditions. In addition, detailed informations about the onset, duration, intensity of the symptoms, and radiological details of CT scans were limited or not obtained in our retrospective analysis.

In conclusion, this preliminary case series does not support an excess morbidity and mortality among symptomatic COVID-19 PLWH and with viral suppression on ART. SARS-CoV-2 infections may occur during boosted darunavir-based and/or on tenofovir containing ART. Larger studies are needed to elucidate any protective or deleterious effect of HIV and antiretroviral therapy.

## Data Availability

The datasets generated during and/or analysed during the current study are available from the corresponding author on reasonable request.

## References

1. World Health Organization (WHO): Coronavirus disease (COVID-19) outbreak situation. https://www.who.int/emergencies/diseases/novel-coronavirus-2019

2. Guan WJ, Ni ZY, Hu Y, et al. Clinical Characteristics of Coronavirus Disease 2019 in China. N Engl J Med. 2020 Feb 28. https://doi.org/10.1056/NEJMoa2002032

3. Liang W, Guan W, Chen R, et al. Cancer patients in SARS-CoV-2 infection: a nationwide analysis in China. Lancet Oncol. 2020; 21: 335–337.

4. Shi Y, Yu X, Zhao H, Wang H, Zhao R, Sheng J. Host susceptibility to severe COVID-19 and establishment of a host risk score: findings of 487 cases outside Wuhan. Crit Care. 2020;24:108.

5. Zhou F, Yu T, Du R, et al. Clinical course and risk factors for mortality of adult inpatients with COVID-19 in Wuhan, China: a retrospective cohort study. Lancet. 2020 Mar 28;395(10229):1054–1062. doi: 10.1016/S0140-6736(20)30566-3. Epub 2020 Mar 11.

6. Blanco JL, Ambrosioni J, Garcia F, Martínez E, Soriano A, Mallolas J, Miro JM; COVID-19 in HIV Investigators. COVID-19 in patients with HIV: clinical case series. Lancet HIV. 2020 Apr 15. pii: S2352–3018(20)30111–9. doi: 10.1016/S2352-3018(20)30111-9. [Epub ahead of print

7. He ZC, Dong Q, Zhuang H, Song S, Peng G, Dwyer DE. Effects of severe acute respiratory syndrome (SARS) coronavirus infection on peripheral blood lymphocytes and their subsets. Int J Infect Dis. 2005;9:323–30.

8. Chen G, Wu D, Guo W, et al. Clinical and immunologic features in severe and moderate Coronavirus Disease 2019. J Clin Invest. 2020 Mar 27. pii: 137244.

9. Qin C, Zhou L, Hu Z, et al. Dysregulation of immune response in patients with COVID-19 in Wuhan, China [published online ahead of print, 2020 Mar 12]. Clin Infect Dis. 2020; doi:10.1093/cid/ciaa248

10. Wu Z, McGoogan JM. Characteristics of and Important Lessons from the Coronavirus Disease 2019 (COVID-19) Outbreak in China: Summary of a Report of 72-314 Cases from the Chinese Center for Disease Control and Prevention. JAMA. 2020 Feb 24. doi: 10.1001/jama.2020.2648.

11. Peters PJ, Skarbinski J, Louie JK, et al. HIV-infected hospitalized patients with 2009 pandemic influenza A (pH1N1)—United States, spring and summer 2009. Clin Infect Dis. 2011; 52 Suppl 1:S183–8.

12. Lynfield R, Davey R, Dwyer DE, et al. Outcomes of influenza A (H1N1) pdm09 virus infection: results from two international cohort studies. PLoS One. 2014; 9(7):e101785. doi: 10.1371/journal.pone.0101785.

13. Wong AT, Tsang OT, Wong MY, et al. Coronavirus infection in an AIDS patient. AIDS. 2004; 18: 829–30.

14. Zhu F, Cao Y, Xu S, Zhou M. Co-infection of SARS-CoV-2 and HIV in a patient in Wuhan city, China [published online March 11, 2020]. J Med Virol. https://doi.org/10.1002/jmv.25732

15. EACS & BHIVA Statement on risk of COVID-19 for people living with HIV (PLWH). https://www.eacsociety.org/home/covid-19-and-hiv.html

16. Robert-Koch Institute, Berlin. Germany. Corona Virus Disease 2019. Daily Situation Report. Last access date: 25th April 2020. https://www.rki.de/DE/Content/InfAZ/N/Neuartiges_Coronavirus/Situationsberichte/2020-04-25-en.pdf?__blob=publicationFile

17. Richardson S, Hirsch JS, Narasimhan M, et al. Presenting Characteristics, Comorbidities, and Outcomes Among 5700 Patients Hospitalized With COVID-19 in the New York City Area. JAMA. 2020 Apr 22. doi: 10.1001/jama.2020.6775. [Epub ahead of print]

18. Mizumoto K, Kagaya K, Zarebski A, Chowell G. Estimating the asymptomatic proportion of coronavirus disease 2019 (COVID-19) cases on board the Diamond Princess cruise ship, Yokohama, Japan, 2020. Euro Surveill. 2020 Mar;25(10). doi: 10.2807/1560-7917.ES.2020.25.10.2000180.

19. Gudbjartsson DF, Helgason A, Jonsson H, et al. Spread of SARS-CoV-2 in the Icelandic Population. N Engl J Med. 2020 Apr 14. doi: 10.1056/NEJMoa2006100. [Epub ahead of print]

20. Stecher M, Schommers P, Schmidt D, et al. Antiretroviral treatment indications and adherence to the German-Austrian treatment initiation guidelines in the German ClinSurv HIV Cohort between 1999 and 2016. Infection. 2019; 47: 247–255.

21. Machnowska P, Meixenberger K, Schmidt D, et al.: Prevalence and persistence of transmitted drug resistance mutations in the German HIV-1 Seroconverter Study Cohort. PLoS ONE 2019;14(1): e0209605. https://doi.org/10.1371/journal.pone.0209605

22. Lim J, Jeon S, Shin HY, et al. Case of the Index Patient Who Caused Tertiary Transmission of COVID-19 Infection in Korea: the Application of Lopinavir/Ritonavir for the Treatment of COVID-19 Infected Pneumonia Monitored by Quantitative RT-PCR. J Korean Med Sci. 2020; 35:e79.

23. Wang Z, Chen X, Lu Y, Chen F, Zhang W. Clinical characteristics and therapeutic procedure for four cases with 2019 novel coronavirus pneumonia receiving combined Chinese and Western medicine treatment. Biosci Trends. 2020; 14: 64–68.

24. Wu J, Li W, Shi X, et al. Early antiviral treatment contributes to alleviate the severity and improve the prognosis of patients with novel coronavirus disease (COVID-19). J Intern Med. 2020 Mar 27. https://doi.org/10.1111/joim.13063

25. Park SY, Lee JS, Son JS, et al. Post-exposure prophylaxis for Middle East respiratory syndrome in healthcare workers. J Hosp Infect. 2019; 101: 42–46.

26. Cao B, Wang Y, Wen D, et al. A Trial of Lopinavir-Ritonavir in Adults Hospitalized with Severe Covid-19. N Engl J Med. 2020 Mar 18. https://doi.org/10.1056/NEJMoa2001282

27. U.S. Department of Health and Human Services. Interim Guidance for COVID-19 and Persons with HIV. https://aidsinfo.nih.gov/guidelines/html/8/covid-19-and-persons-with-hiv--interim-guidance-/554/interim-guidance-for-covid-19-and-persons-with-hiv.

28. Sanders JM, Monogue ML, Jodlowski TZ, Cutrell JB. Pharmacologic Treatments for Coronavirus Disease 2019 (COVID-19): A Review. JAMA. 2020 Apr 13. doi: 10.1001/jama.2020.6019.

29. Elfiky AA. Ribavirin, Remdesivir, Sofosbuvir, Galidesivir, and Tenofovir against SARS-CoV-2 RNA dependent RNA polymerase (RdRp): A molecular docking study. Life Sci. 2020 Mar 25: 117592.

30. Randomized Clinical Trial for the Prevention of SARS-CoV-2 Infection (COVID-19) in Healthcare Personnel (EPICOS). https://clinicaltrials.gov/ct2/show/NCT04334928?cond=Randomized+clinical+trial+for+the+prevention+of+SARS-CoV-2+infection+%28COVID-19%29+in+healthcare+personnel&draw=2&rank=1

